# Prognostic Impact of Hemodynamic Transitions in Pulmonary Artery Pulsatility Index and Compliance in Advanced Heart Failure: A Post-hoc Analysis of the ESCAPE Trial

**DOI:** 10.1101/2025.10.03.25337300

**Authors:** Yuta Ozaki, Yusuke Uemura, Toru Kondo, Shingo Kazama, Shogo Yamaguchi, Takashi Okajima, Takayuki Mitsuda, Shinji Ishikawa, Kenji Takemoto, Takahiro Okumura, Masato Watarai, Toyoaki Murohara

## Abstract

**Background:** Advanced heart failure (HF) remains associated with poor outcomes despite contemporary therapies. Right ventricular (RV) dysfunction, a hallmark of advanced HF, is strongly influenced by afterload. We aimed to evaluate whether the combined assessment of the pulmonary artery pulsatility index (PAPi) and pulmonary arterial capacitance (PAC)-and their transitions during acute-phase therapy-provides prognostic stratification in patients with advanced HF.

**Methods:** We conducted a post-hoc analysis of the Evaluation Study of Congestive Heart Failure and Pulmonary Artery Catheterization Effectiveness (ESCAPE) trial, including 146 patients with complete hemodynamic data. A bootstrap-based grid search identified prognostic cutoffs: PAPi at 2.67 and PAC at 2.03. The *optimal* zone was defined as PAPi ≥2.67 and PAC ≥2.03; the *suboptimal* zone as PAPi <2.67 or PAC <2.03. Patients were categorized into four groups based on transitions between zones from baseline to the final assessment after acute-phase therapy. The primary endpoint was a composite of all-cause mortality, left ventricular assist device implantation, or heart transplantation within 6 months.

**Results:** At baseline, 127 patients were in the *suboptimal* zone. Following acute-phase therapy, 33 transitioned to the *optimal* zone, while 94 remained *suboptimal*. Kaplan–Meier curves demonstrated significant stratification among groups. In Cox regression models using the *suboptimal→suboptimal* as reference, transition to the *suboptimal→optimal* was associated with improved prognosis (multivariable hazard ratio 0.300, 95% confidence interval 0.107–0.847, P=0.023).

**Conclusions:** Transitions in PAPi and PAC during acute-phase therapy were associated with subsequent outcomes in advanced HF. Combined assessment of PAPi and PAC may provide a novel therapeutic target for risk stratification and management in this high-risk population.

## Introduction

Advanced heart failure (HF) remains associated with poor prognosis despite contemporary therapies (1). Right ventricular (RV) dysfunction is not only a hallmark of advanced HF but also an established predictor of adverse outcomes (2). However, limited evidence is available regarding how dynamic changes in RV function during acute decompensation and subsequent treatment affect prognosis in this high-risk population.

RV function is highly sensitive to afterload, underscoring the need to assess both contractile performance and loading conditions simultaneously (3–5). The pulmonary artery pulsatility index (PAPi), calculated as pulmonary artery pulse pressure divided by right atrial pressure, has emerged as a robust marker of RV function and has been shown to predict adverse outcomes in advanced HF (6–8). Pulmonary arterial capacitance (PAC), defined as stroke volume divided by pulmonary artery pulse pressure, reflects the pulsatile component of RV afterload and is particularly responsive to elevations in left-sided filling pressures (9). We previously demonstrated that the combined assessment of PAPi and PAC stratifies risk and identifies distinct phenotypes in patients with compensated HF with preserved ejection fraction (HFpEF) and HF with reduced ejection fraction (HFrEF) (10, 11).

Building on these observations, we aimed to determine whether transitions in PAPi and PAC in response to acute-phase therapy are associated with subsequent clinical outcomes in patients with advanced HF. To address this question, we analyzed data from the North American Evaluation Study of Congestive Heart Failure and Pulmonary Artery Catheterization Effectiveness (ESCAPE) trial, which uniquely provides serial invasive hemodynamic assessments during hospitalization for decompensated advanced HF (12).

## Methods

### Study Population

We conducted a post hoc analysis of de-identified, individual-level data from the ESCAPE trial, accessed through the Biologic Specimen and Data Repository Information Coordinating Center (BioLINCC) of the United States National Heart, Lung, and Blood Institute (NHLBI). Before data access, a signed data use agreement was completed. The study was exempt from institutional review board oversight, as determined by the Anjo Kosei Hospital ethics committee (Waiver No. R24-031).

The ESCAPE trial enrolled 433 patients hospitalized with advanced HF across 26 centers in the United States and Canada between January 2000 and November 2003. The trial’s design and principal results have been described previously (12). In summary, patients with severe, symptomatic HF were randomized to either therapy guided by pulmonary artery catheterization or by clinical assessment alone. Eligibility criteria included a left ventricular ejection fraction ≤30%, persistent New York Heart Association class IV symptoms, at least one hospitalization for decompensated HF within the previous 6 months, and a documented history of HF of at least 3 months. Additional requirements included ongoing treatment with angiotensin-converting enzyme inhibitors and diuretics for at least 3 months before randomization, a systolic blood pressure ≤125 mmHg, and clinical signs of elevated filling pressures. Major exclusion criteria included serum creatinine level >3.5 mg/dL and recent or ongoing use of high-dose inotropes. Patients assigned to the pulmonary artery catheterization-guided arm were managed to achieve hemodynamic targets-specifically, pulmonary capillary wedge pressure ≤15 mmHg and right atrial pressure ≤8 mmHg.- alongside clinical objectives. Pulmonary artery catheters were typically maintained until target hemodynamic goals were reached and a discharge vasodilator regimen had been established, in accordance with study protocol. The use of intravenous inotropes was discouraged throughout hospitalization.

For this analysis, we included patients with complete invasive hemodynamic data required to calculate both PAPi and PAC. Two distinct time points were analyzed: (1) baseline values obtained immediately after pulmonary artery catheter insertion at the time of randomization, and (2) final measurements recorded just prior to catheter removal following acute-phase therapy. PAPi was calculated as pulmonary artery pulse pressure (systolic minus diastolic pulmonary artery pressure) divided by right atrial pressure, and PAC was defined as stroke volume divided by pulmonary artery pulse pressure. Because stroke volume was not directly measured, it was estimated as cardiac output divided by heart rate.

The primary study endpoint was a composite of all-cause mortality, orthotopic heart transplantation, or left ventricular assist device (LVAD) implantation within 6 months of randomization.

### Statistical analysis

Continuous variables were summarized as mean ± standard deviation or median with interquartile range (IQR), depending on their distribution as assessed by the Shapiro-Wilk test. Categorical variables were reported as counts and percentages. Paired comparisons between baseline and final hemodynamic values were performed using either paired t-tests or Wilcoxon signed-rank tests, based on variable distribution.

To identify prognostically relevant thresholds for PAPi and PAC, we applied a grid search to the final hemodynamic data obtained at catheter removal. Each variable was dichotomized at 10-percentile intervals between the 30th and 70th percentiles, and all possible cutoff combinations were tested in Cox proportional hazards models. Model performance was assessed using the concordance index (C-index), and the threshold pair that consistently yielded the highest C-index across 10,000 bootstrap iterations was selected. This process identified PAPi 2.03 and PAC 2.67 as the optimal thresholds.

Patients were categorized at both baseline and final assessments into one of two hemodynamic zones:

- “*optimal*” zone: PAPi ≥2.03 and PAC ≥2.67,
- “*suboptimal*” zone: either PAPi <2.03 or PAC <2.67.

Transitions between zones from baseline to final assessment were classified into four groups:

1. *optimal → optimal,*
2. *optimal → suboptimal,*
3. *suboptimal → optimal,*
4. *suboptimal → suboptimal*.

Kaplan–Meier survival curves were generated for both the dichotomized hemodynamic zones and the four transition categories. Cox proportional hazards models were used to estimate hazard ratios (HRs) and 95% confidence intervals (CIs). Multivariable Cox models were adjusted for the validated ESCAPE discharge score (13). The discharge score, derived from the ESCAPE dataset, includes the following predictors assessed at discharge: age >70 years, blood urea nitrogen (BUN) >40 mg/dL, BUN >90 mg/dL, 6-minute walk distance <300 feet, serum sodium <130 mmol/L, loop diuretic dose >240 mg/day, absence of β-blocker use, B-type natriuretic peptide (BNP) >500 pg/mL (each assigned 1 point), BNP >1300 pg/mL (+3 points), and for in-hospital cardiac arrest or mechanical ventilation (+2 points), yielding a maximum possible score of 13 (13). Missing values for individual score components were imputed using multiple imputation.

The proportions of patients who received loop diuretics, intravenous vasodilators, and intravenous inotropes (milrinone, dopamine, or dobutamine) during hospitalization were compared across the four transition groups. Group-level differences were assessed using χ² tests, with a significance threshold of P <0.05.

All statistical analyses were performed using R version 4.3.2 (R Foundation for Statistical Computing, Vienna, Austria). The following R packages were used: “*survival*”, “*survminer*”, “*purr*”, and *“mice”*. A two-sided P value <0.05 was considered statistically significant.

## Results

Among the 433 participants in the ESCAPE trial, invasive hemodynamic data were available for 202 patients, including 187 in the pulmonary artery catheter-guided therapy arm and 15 in the standard care arm who underwent right heart catheterization. Of these, 146 patients had complete baseline data for both PAPi and PAC at enrollment and at the final assessment before pulmonary artery catheter removal, and were included in the present analysis cohort.

Table 1 summarizes baseline and pre-discharge patient characteristics. The mean age was 56.2 years, 39 patients (26.9%) were female, and the median left ventricular ejection fraction was 20.0%. At discharge, the median BNP level was 395.5 pg/mL, and the median discharge score was 2.0 (IQR, 1.0–4.0) points.

**Table 1.** Baseline and pre-discharge clinical characteristics. Demographics, comorbidities, laboratory values, medications, and discharge score are reported. Data are summarized as medians (interquartile ranges), means ± standard deviations, or n (%). ACE, angiotensin-converting enzyme; ARBs, angiotensin receptor blockers.

| Characteristics | Baseline | Pre-discharge |
| --- | --- | --- |
| **Demographics** |  |  |
| Age (years) | 56.2 ± 13.9 | — |
| Female | 39 (26.9%) | — |
| Race |  | — |
| White | 83 (58.5%) | — |
| Black | 44 (31.0%) | — |
| Asian | 0 (0.0%) | — |
| Other | 15 (10.6%) | — |
| Body mass index (kg/m<sup>2</sup>) | 27.7 (23.4, 33.1) | — |
| **Comorbidities** | 20.0 (15.0, 23.0) | — |
| Ischemic heart disease | 79 (54.1%) | — |
| Valvular heart disease | 8 (5.5%) | — |
| Hypertension | 72 (49.3%) | — |
| Diabetes | 47 (33.3%) | — |
| Atrial fibrillation | 45 (30.8%) | — |
| Chronic obstructive pulmonary disease | 24 (16.4%) | — |
| **Echocardiographic finding** |  |  |
| Ejection fraction (%) | 20.0 (15.0, 23.0) | — |
| **Laboratory findings** |  |  |
| Sodium (mmol/L) | 137.0 (135.0, 140.0) | 135.0 (133.0, 138.0) |
| Blood urea nitrogen (mg/dL) | 29.0 (20.0, 45.2) | 32.0 (21.8, 49.0) |
| Creatinine (mg/dL) | 1.40 (1.10, 1.80) | 1.50 (1.10, 1.90) |
| Hemoglobin (mg/dL) | 12.3 ± 1.7 | 11.7 (10.7, 13.5) |
| B-type natriuretic peptide (pg/mL) | 565.5 (249.0, 1,303.5) | 395.5 (174.0, 855.8) |
| **Medications** |  |  |
| ACE inhibitors | 114 (78.1%) | 109 (75.2%) |
| ARBs | 30 (20.5%) | 16 (11.0%) |
| Beta-blockers | 92 (63.0%) | 77 (53.5%) |
| Diuretics | 143 (97.9%) | 132 (91.0%) |
| Isosorbide mononitrate | 27 (18.5%) | — |
| Isosorbide dinitrate | 19 (13.0%) | — |
| Dobutamine | 14 (9.9%) | — |
| Dopamine | 4 (2.9%) | — |
| Milrinone | 2 (1.4%) | — |
| **Discharge score** | – | 2.0 (1.0, 4.0) |

Table 2 presents invasive hemodynamic parameters at baseline and at the final assessment prior to pulmonary artery catheter removal. Following acute-phase therapy, filling pressures declined, and cardiac output increased. Notably, both PAPi and PAC improved significantly post-treatment, with median values rising from 2.37 to 3.00 and from 1.66 to 2.37, respectively.

**Table 2.**
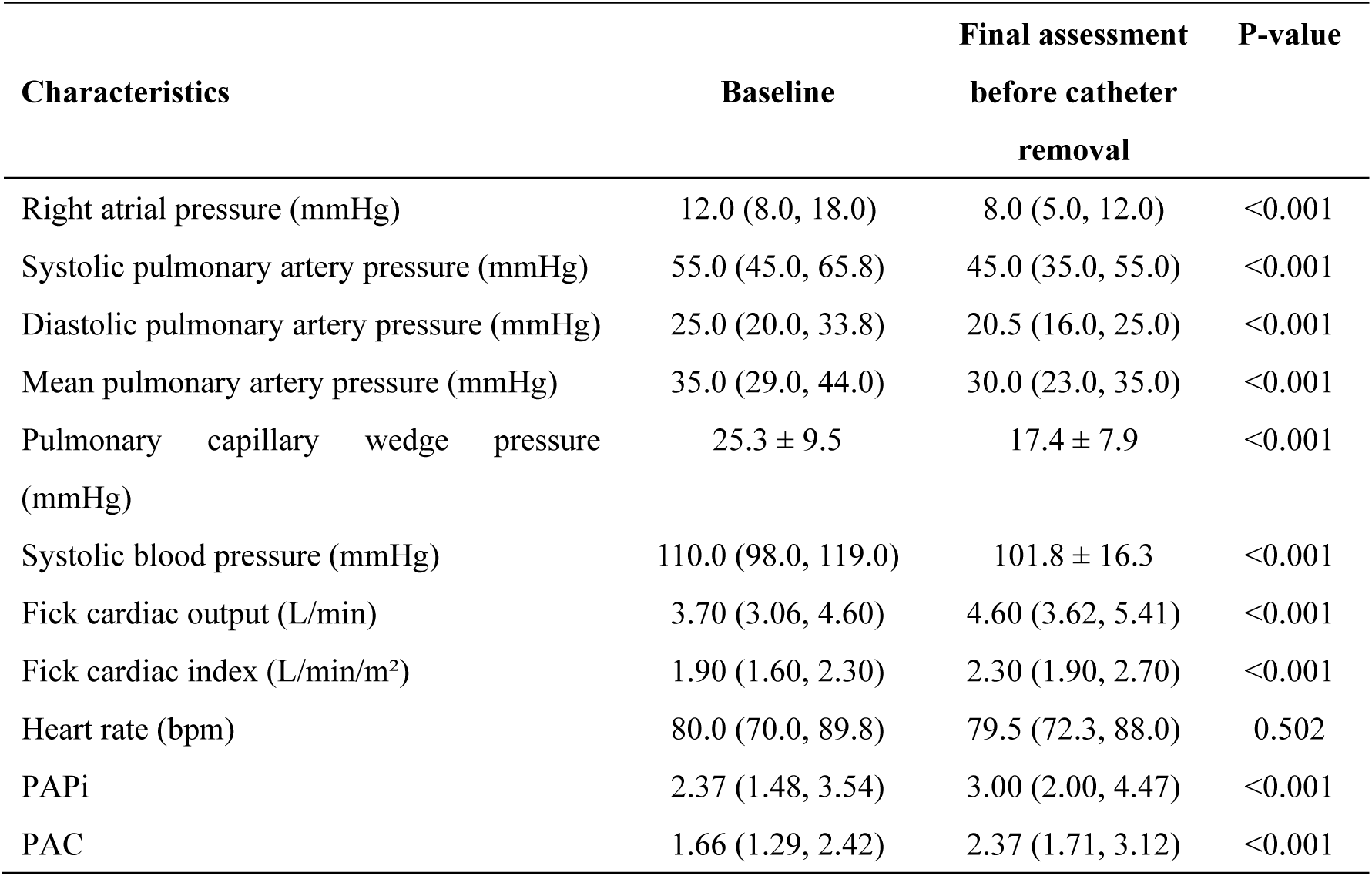
Hemodynamics at baseline and at the final assessment. Invasive hemodynamic parameters were measured at baseline and at the final assessment prior to pulmonary artery catheter removal. Values are summarized as medians (interquartile ranges) or means ± standard deviations. Paired comparisons were performed using paired t-tests or Wilcoxon signed-rank tests, depending on data distribution. PAC, pulmonary arterial capacitance; PAPi, pulmonary artery pulsatility index.

To determine the prognostically relevant thresholds of PAPi and PAC at the final assessment, we conducted a grid search with bootstrap resampling. Across 10,000 iterations, the combination of PAPi 2.67 and PAC 2.03 most consistently emerged as the optimal cutoff, appearing in 126 runs with a mean C-index of 0.699. This combination was therefore selected as the threshold for outcome prediction (Figure 1). Based on these thresholds, patients were stratified into four groups according to PAPi and PAC status at final assessment. Kaplan– Meier analysis demonstrated clear separation of survival curves, with significantly different outcomes across strata (log-rank P = 0.005) (Figure 2).

**Figure 1.**
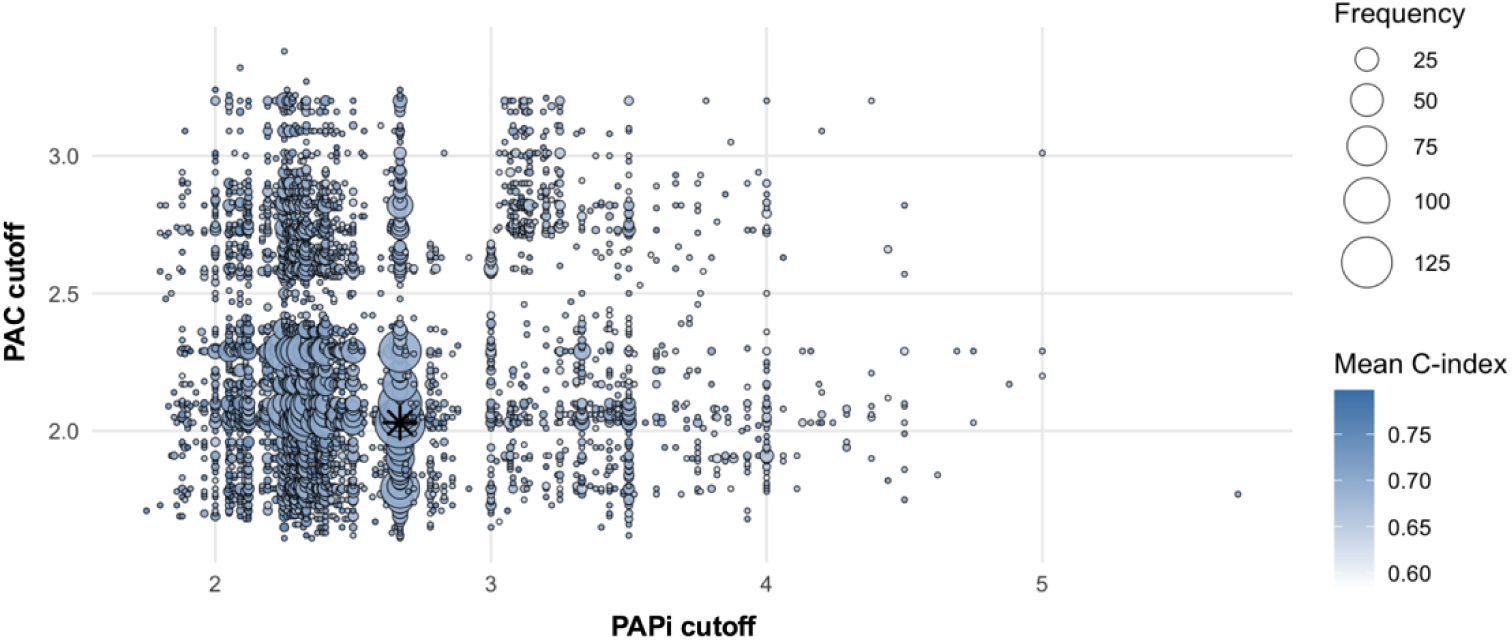
Grid search with bootstrap resampling to define prognostic cutoffs for PAPi and PAC. A bootstrap-based grid search was conducted using the final hemodynamic data at the time of pulmonary artery catheter removal to identify prognostic cutoffs for PAPi and PAC. Each variable was dichotomized at 10-percentile intervals between the 30th and 70th percentiles, and all combinations were tested using Cox proportional hazards models across 10,000 bootstrap iterations. The combination of PAPi 2.67 and PAC 2.03 most consistently produced the highest concordance index (C-index), denoted by the star marker. PAC, pulmonary arterial capacitance; PAPi, pulmonary artery pulsatility index.

**Figure 2.**
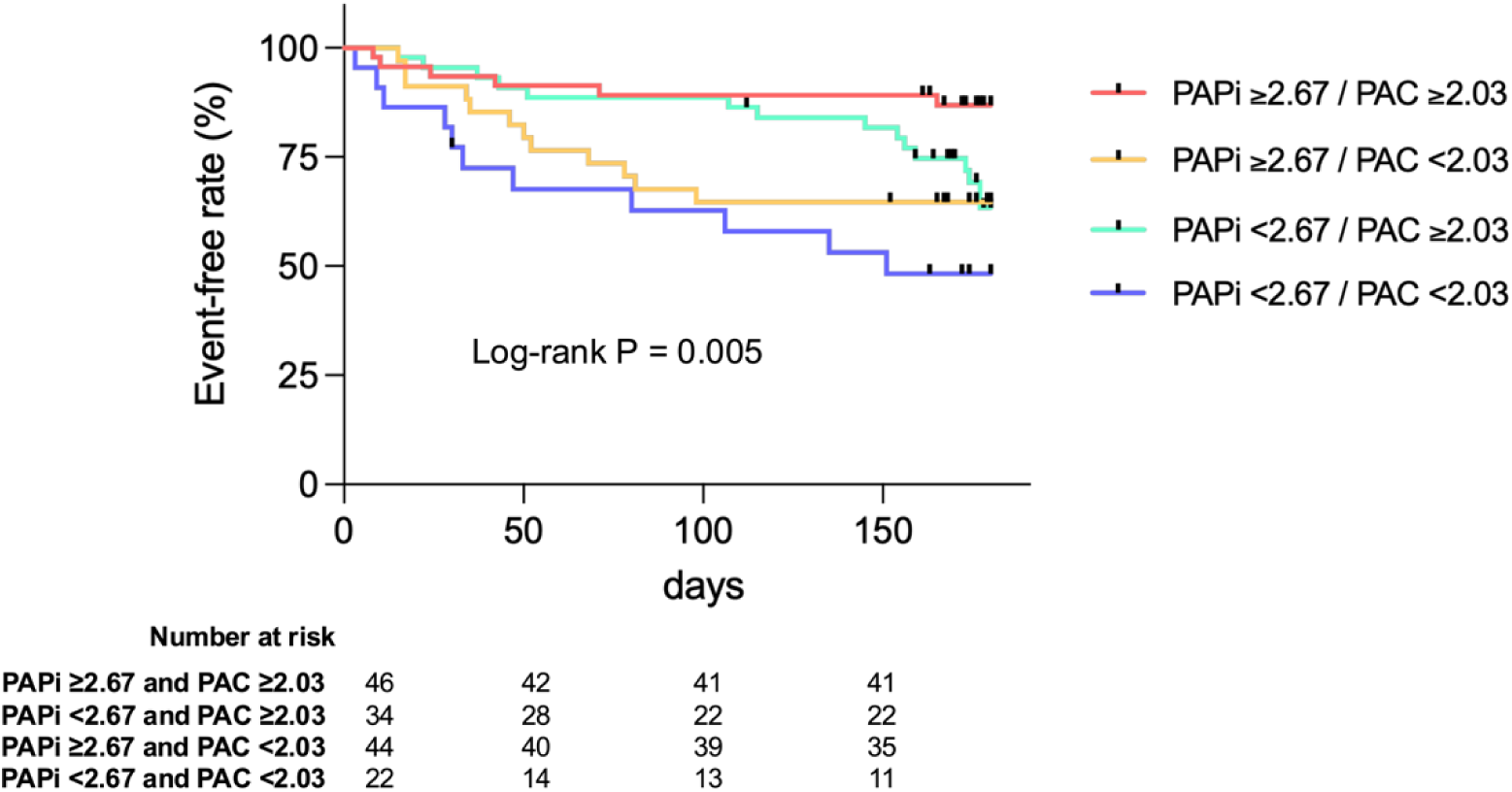
Kaplan–Meier curves stratified by PAPi and PAC at the final assessment. Kaplan–Meier curves for the composite endpoint of all-cause death, left ventricular assist device implantation, or heart transplantation within 6 months, stratified by four strata according to the prognostic cutoffs of PAPi (2.67) and PAC (2.03) at the final assessment. PAC, pulmonary arterial capacitance; PAPi, pulmonary artery pulsatility index.

We next defined the stratum with PAPi ≥2.67 and PAC ≥2.03 as the *optimal* zone, and any value of PAPi <2.67 or PAC <2.03 as the *suboptimal* zone. Patients in the *suboptimal* zone at the time of pulmonary artery catheter removal had poorer outcomes than those in the *optimal* zone (log-rank P = 0.006; Supplemental Figure 1; multivariable Cox regression adjusted for the discharge score: hazard ratio, 2.620; 95% CI; 1.100-6.245; P = 0.030; Supplemental Table 1). Based on transitions from baseline to final assessment, patients were categorized into four groups: *optimal→optimal* (n=13), *optimal→suboptimal* (n=6), *suboptimal→optimal* (n=33), and *suboptimal→suboptimal* (n=94). Kaplan–Meier curves demonstrated significant stratification of outcomes across these groups, with the *suboptimal→suboptimal* group showing the poorest prognosis (log-rank P = 0.016) (Figure 3). In Cox proportional hazards analyses using the *suboptimal→suboptimal* group as the reference, the *suboptimal→optimal* group had significantly better prognosis. The hazard ratios were 0.262 (95% CI 0.093–0.737, P = 0.011) in the univariable model, and 0.300 (95% CI 0.107–0.847, P = 0.023) in the multivariable model (adjusted for the discharge score). In contrast, the *optimal→optimal* and *optimal→suboptimal* groups did not differ significantly from the reference group (Table 3).

**Figure 3.**
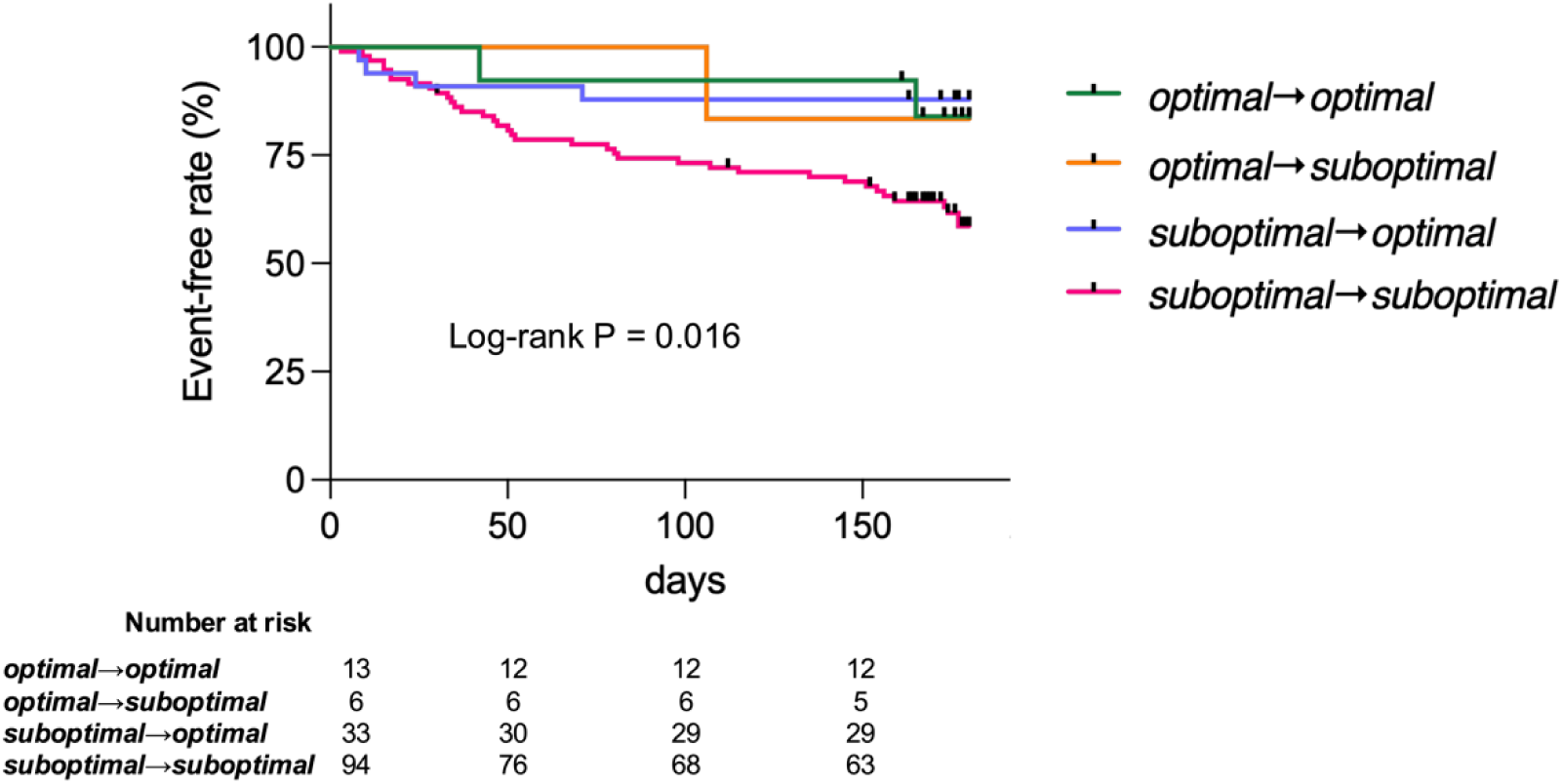
Kaplan–Meier curves according to transitions in PAPi and PAC from baseline to final assessment. The *optimal* zone was defined as PAPi ≥2.67 and PAC ≥2.03, and the *suboptimal* zone as PAPi <2.67 or PAC <2.03. Patients were classified into four groups according to transitions between baseline and the final assessment: *optimal→optimal* (n=13), *optimal→suboptimal* (n=6), *suboptimal→optimal* (n=33), and *suboptimal→suboptimal* (n=94). Kaplan–Meier curves for the composite endpoint of all-cause death, left ventricular assist device implantation, or heart transplantation within 6 months are shown, stratified by these groups. PAC, pulmonary arterial capacitance; PAPi, pulmonary artery pulsatility index.

**Table 3.** Associations between transitions in PAPi and PAC and the composite endpoint. Cox proportional hazards models for the composite endpoint of all-cause death, left ventricular assist device implantation, or heart transplantation within 6 months. The *optimal* zone was defined as PAPi ≥2.67 and PAC ≥2.03, the *suboptimal* zone as PAPi <2.67 or PAC <2.03. Patients were categorized into four groups according to transitions between baseline and the final assessment: *optimal→optimal* (n=13), *optimal→suboptimal* (n=6), *suboptimal→optimal* (n=33), and *suboptimal→suboptimal* (n=94). Hazard ratios with 95% CIs are reported for the univariate and multivariable models adjusted for the validated discharge score. CI, confidence interval; PAC, pulmonary arterial capacitance; PAPi, pulmonary artery pulsatility index.

|  | Univariable |  | Multivariable |  |
| --- | --- | --- | --- | --- |
|  | Hazard ratio<br>(95% CI) | P-value | Hazard ratio<br>(95% CI) | P-value |
| *Optimal* → *Optimal* | 0.328<br>(0.079–1.360) | 0.125 | 0.740<br>(0.169–3.240) | 0.689 |
| *Optimal* → *Suboptimal* | 0.341<br>(0.047–2.490) | 0.289 | 0.523<br>(0.071–3.860) | 0.525 |
| *Suboptimal* → *Optimal* | 0.262<br>(0.093–0.737) | 0.011 | 0.300<br>(0.107–0.847) | 0.023 |
| *Suboptimal* → *Suboptimal* (Ref) | 1.00 | – | 1.00 | – |

The proportions of patients receiving loop diuretics, intravenous vasodilators, and intravenous inotropes (milrinone, dopamine, or dobutamine) during hospitalization were compared across the four transition groups. Among these, only vasodilator use differed significantly: the *suboptimal→optimal* and *suboptimal→suboptimal* groups had higher utilization rates compared with the other groups. No statistically significant differences were observed in the use of loop diuretics or inotropes across the groups (Supplemental Figure 2).

## Discussion

In this post-hoc analysis of the ESCAPE trial, we identified two principal findings. First, the combination of PAPi and PAC measured at the final hemodynamic assessment following acute-phase therapy effectively stratified prognosis in patients with advanced HF. Second, beyond these cross-sectional measurements, transitions in PAPi and PAC from baseline to final assessment provided additional prognostic information, underscoring the incremental value of incorporating temporal changes in RV hemodynamics into risk assessment.

Despite contemporary therapies, the prognosis of advanced HF remains poor (1). RV function is a critical determinant of adverse outcomes in this population (2). Because RV performance is strongly influenced by afterload, comprehensive evaluation requires simultaneous assessment of both contractile function and loading conditions (3–5). The concept of RV-pulmonary arterial (RV–PA) coupling integrates these elements, and echocardiographic surrogates of RV–PA coupling have been shown to stratify outcomes (14, 15). However, echocardiographic assessment of RV function has intrinsic limitations, and accurate quantification of RV–PA coupling requires pressure–volume loop analysis, which is not feasible in routine clinical practice. Recently, PAPi has been reported as a marker of RV function with prognostic value (6–8). Building on this, we examined the combined use of PAPi and PAC, the latter reflecting the pulsatile component of RV afterload. This approach extends our previous observations in compensated HFpEF and HFrEF to patients with advanced HF (10, 11).

A distinctive aspect of this study is the analysis of transitions in PAPi and PAC during acute-phase therapy, through which we identified a hemodynamic target zone, defined as the *optimal* zone (PAPi ≥2.67 and PAC ≥2.03). At baseline, 87.0% of patients were classified in the *suboptimal* zone. Following acute-phase therapy, 22.6% improved to the *optimal* zone, whereas 64.4% remained in the *suboptimal* zone. Patients who persisted in the *suboptimal* zone had the poorest outcomes, while those who improved demonstrated a more favorable prognosis. Collectively, these findings suggest that PAPi and PAC may serve as therapeutic targets in the management of advanced HF.

Therapeutic strategies to improve the combined status of PAPi and PAC remain undefined. In this study, we examined in-hospital medication use captured in the ESCAPE dataset; however, no consistent associations with hemodynamic improvement were identified. This highlights the need for therapeutic approaches beyond conventional treatments. Agents unavailable at the time of the ESCAPE trial, such as angiotensin receptor–neprilysin inhibitors and sodium–glucose cotransporter-2 inhibitors, have been reported to improve RV– PA coupling and may represent promising treatment options (16, 17). In addition, emerging agents such as omecamtiv mecarbil and levosimendan may provide benefit in selected patients (18, 19). For patients who fail to achieve improvement despite these interventions, timely referral for advanced therapies such as mechanical circulatory support or heart transplantation should be considered. Further studies are warranted to establish effective interventions targeting PAPi and PAC as modifiable treatment goals in advanced HF.

Several limitations of this study should be acknowledged. First, although our analysis was based on the prospective ESCAPE trial, it was post-hoc and therefore subject to inherent risks of bias. Second, because ESCAPE enrolled only patients with advanced HF, the generalizability of our findings-including the *optimal* zone thresholds for PAPi (≥2.67) and PAC (≥2.03), is uncertain and requires external validation in independent cohorts. Third, the dataset did not capture detailed temporal trajectories of acute-phase therapies, precluding more granular analyses of treatment effects. Finally, as the ESCAPE trial was conducted between 2000 and 2003, current management strategies for acute advanced HF have since evolved. Nonetheless, even with contemporary therapies, the prognosis of advanced HF remains poor, underscoring the need for refined risk stratification and novel therapeutic targets.

In summary, transitions in PAPi and PAC during acute-phase therapy were closely associated with subsequent outcomes in patients with advanced HF. Assessment based on the combined status of PAPi and PAC may represent a novel therapeutic target for risk stratification and management in this high-risk population, whose prognosis remains poor despite advances in contemporary therapy.

## Data Availability

The data underlying this article are available from the National Heart, Lung, and Blood Institute (NHLBI) BioLINCC repository. Access to the ESCAPE trial dataset can be obtained upon submission and approval of a data use request at https://biolincc.nhlbi.nih.gov/studies/escape/.

## Acknowledgments

The authors acknowledge the National Heart, Lung, and Blood Institute (NHLBI) and the Biologic Specimen and Data Repository Information Coordinating Center (BioLINCC) for providing access to the ESCAPE trial dataset. The data used in this study were obtained from BioLINCC for this analysis under an approved data request. The authors are solely responsible for the design, analysis, interpretation, and conclusions of this study, which do not necessarily reflect the views of the NHLBI or BioLINCC.

## Supplemental Material

Table S1 Figure S1-S2

## Non-standard Abbreviations and Acronyms

BNP: B-type Natriuretic Peptide
ESCAPE: Evaluation Study of Congestive Heart Failure and Pulmonary Artery Catheterization Effectiveness
HFpEF: Heart Failure with Preserved Ejection Fraction
HFrEF: Heart Failure with Reduced Ejection Fraction
NHLBI: National Heart, Lung, and Blood Institute
PAC: Pulmonary Arterial Capacitance
PAPi: Pulmonary Artery Pulsatility Index
RV–PA: Right Ventricular–Pulmonary Arterial (Coupling)

